# Impact of Advanced Maternal Age and In Vitro Fertilization Technology on Obstetric and Perinatal Outcomes

**DOI:** 10.64898/2025.12.03.25341529

**Authors:** M Domínguez-Moreno, B Walsh, J De-Martín-Hernández, Ángel Chimenea, G. Antiñolo, L García-Díaz

## Abstract

**Purpose:** To analyze obstetric and perinatal outcomes of pregnant women with advanced maternal age (AMA) after undergoing in vitro fertilization (IVF) technology.

**Methods:** A single-center prospective observational study was conducted between January-December 2024. All women aged 40 and older who conceived via IVF were included. Participants were monitored monthly by a dedicated team of maternal-fetal medicine specialists who assessed maternal clinical conditions and fetal development.

**Results:** A total of 128 pregnant women were recruited. Nearly half of them were nulliparous (n=60,46.88%) and a total of 71 conceived by oocyte donation (55.47%).

A remarkable proportion of women experienced preterm rupture of membranes (n=11,8.59%), diabetes mellitus (n=6,4.69%) or hypertensive disorders (n=13,10.32%). Cesarean delivery was performed in 63 cases (50.4%).

Regarding neonatal outcomes, the mean birth weight was 3,027grams and the mean umbilical cord pH was 7.29. Eight newborns (6.45%) required admission to intensive care unit. Postnatal comorbidities were identified in 14 infants, with respiratory complications being the most frequent (12/14).

**Conclusions:** Pregnancies in women of AMA following IVF are associated with remarkable obstetric and neonatal risks. Our findings emphasize that despite meticulous, close and personalized monitoring in a maternal-fetal medicine unit specialized in high-risk pregnancies, not all unfavorable outcomes can be prevented.

## 1. Introduction

Over the past three decades, there has been a dramatic worldwide increase in the number of women postponing childbearing until their late 35s, and even into their 40s and beyond^1–8^. In fact, the rate of first births among women aged 35 to 39 increased sevenfold between 1973 and 2018, and fivefold among women aged 40 to 44 between 1985 and 2018^1^.

This phenomenon has been associated with a range of social, economic, and cultural factors^6,9^. Shifts in socioeconomic conditions—such as increased participation in the labor market, professional aspirations, limited workplace policies supporting motherhood and childcare, and the desire for financial independence before becoming a parent—have all been linked to this trend^1,6,10,11^.

Fertility in women begins to decline in the early 30s and decreases more rapidly after the mid to late 30s^12^. Consequently, there has been a growing trend in the use of assisted reproductive technologies (ART)^13^. Advances in ART -such as in vitro fertilization (IVF)- have increased the likelihood of successful pregnancies in older women, thereby making these procedures more common^14^. This technology has effectively extended the reproductive window, leading to a higher incidence of pregnancies in women beyond the typical biological reproductive age^3,9^.

Taken together, these shifts in family structure and reproductive planning have contributed to an increased rate of pregnancies among women of advanced maternal age (AMA). Although the term AMA lacks a universally agreed-upon definition^1^, it generally refers to women who are aged 35 years or older at the time of delivery, marking the later years of the reproductive lifespan^6,10^. However, because aging is a gradual and multifaceted process, establishing a precise cutoff point is challenging, especially given that age-related effects often emerge progressively^15,16^.

Regardless of the cutoff used, AMA is a well-established risk factor for adverse pregnancy outcomes^15,17^. Data from population registries and large cohort studies consistently associate AMA with increased risks of maternal complications, including hypertensive disorders, gestational diabetes, placental abnormalities, and the need for medical intervention not only during pregnancy and delivery but also in the postpartum period—for instance, in the form of postpartum depression^3,5,6,9,10,12,17,18^. Beyond the immediate pregnancy, AMA may also negatively affect women’s long-term health, as is the case of future cardiovascular disease^10^.

Moreover, AMA has important implications for neonatal outcomes^1^. Older maternal age is associated with higher rates of miscarriage, chromosomal abnormalities, congenital anomalies, fetal growth restriction, preterm birth, small for gestational age infants, low birthweight, and increased admissions to neonatal intensive care units (NICUs)^1,10,12,16,17^.

Despite the well-documented negative impacts of AMA on maternal and neonatal health, current obstetric guidelines offer limited nuance regarding the effective management of older pregnant women throughout the maternity care continuum^1^. And what is more, the combined effect of AMA and IVF on obstetric and perinatal outcomes remains underexplored.

To address this gap, the present study aims to comprehensively evaluate obstetric and perinatal outcomes in women aged 40 years or older who conceive through IVF. Additionally, we explore potential underlying mechanisms that may explain these outcomes.

## 2. Material and methods

### 2.1. Study design

This single-center prospective observational study was conducted at Virgen del Rocío University Hospital, a tertiary care institution and referral center for complex maternal-fetal pathology. Last year, the hospital provided care for 9,258 patients, with a total of 4,532 births attended.

All women aged 40 years or older who conceived via IVF at our center between January and December 2024 were classified as having AMA and were included in the study. The final analytic sample comprised 128 women. Exclusion criteria included inability to complete appropriate prenatal follow-up or refusal to participate. **Figure 1** shows schematically the selection process and the inclusion and exclusion criteria used.

**Figure 1.**
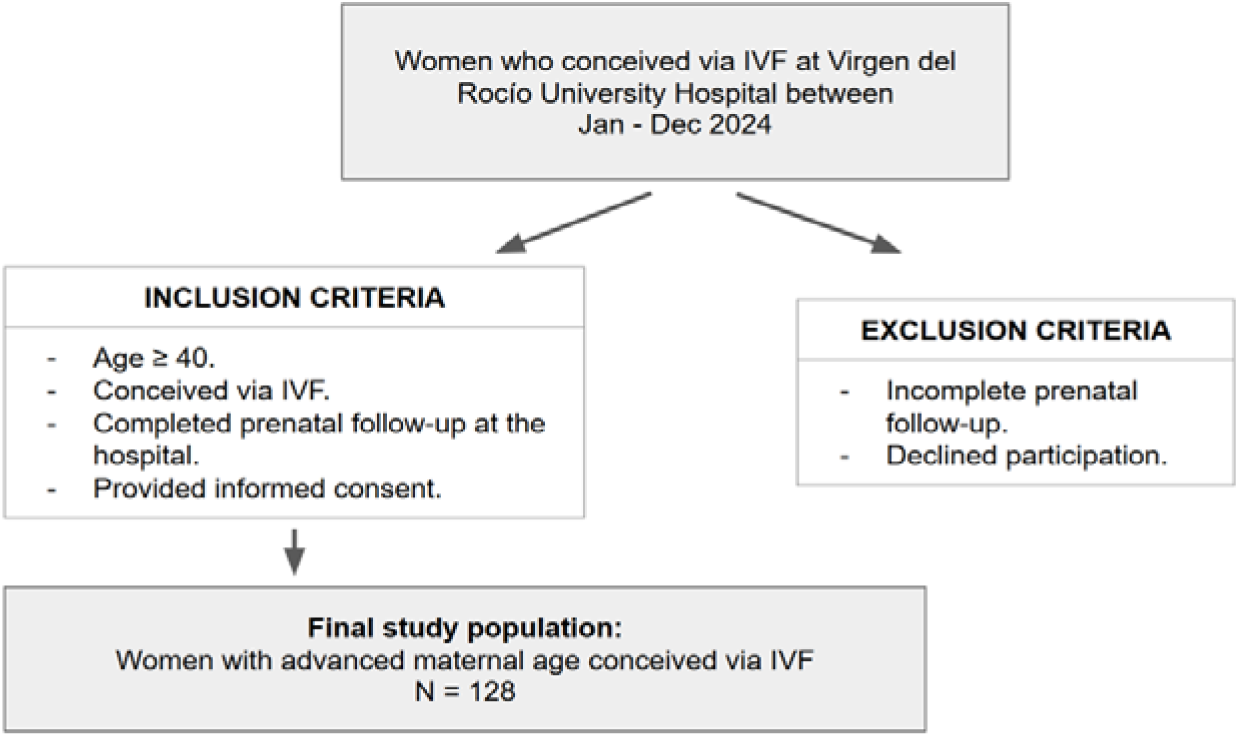
Selection process of the study

Gestational age was determined based on the date of the last menstrual period and confirmed by first-trimester ultrasound. After enrollment, participants were monitored every four to six weeks by a dedicated team of maternal-fetal medicine specialists, with the aim of preventing or detecting obstetric complications. Maternal clinical status was checked including blood pressure reading and fetal development was assessed through well-structured ultrasound examinations, covering biometric parameters and Doppler flowmetry. In addition, serial blood tests were conducted to screen for metabolic disorders and early signs of preeclampsia.

### 2.2. Data collection

Maternal and neonatal characteristics, delivery details, and perinatal complications were documented, encompassing a total of 25 variables. These variables are detailed with their terminology and precise definition in **Table 1**.

**Table 1.** Maternal-fetal variables included in the study.

| Maternal-fetal variables | Definition | Description / value |
| --- | --- | --- |
| <u>Maternal characteristics</u> |  |  |
| Maternal age | Chronological age of the pregnant woman at the time of childbirth. | Continuous variable, expressed in years. |
| Body mass index (BMI) | Calculated as the woman's weight (kg) divided by the square of her height (m<sup>2</sup>). | Continuous variable, expressed in kilograms/metres<sup>2</sup> |
| Weight | Weight of the pregnant woman at the first prenatal visit. | Continuous variable, expressed in kilograms. |
| Height | Height of the pregnant woman at the first prenatal visit. | Continuous variable, expressed in meters. |
| Smoking status | Smoking during pregnancy or within three months prior to conception. | Nominal variable. |
| <u>Obstetric characteristics</u> |  |  |
| Diabetes mellitus | Diagnosis of diabetes confirmed during pregnancy via oral glucose tolerance test or abnormal glycemic profiles. | Nominal variable. |
| Chronic hypertension | Hypertension diagnosed prior to pregnancy or before 20 weeks of gestation. | Nominal variable. |
| Gravity | Total number of prior pregnancies at the time of assessment. | Continuous variable. |
| History of miscarriage | Number of spontaneous pregnancy losses before 23 weeks of gestation. | Continuous variable. |
| Parity | Number of previous deliveries at the time of the assessment. | Continuous variable. |
| Previous cesarean section | History of cesarean delivery in any prior pregnancy. | Continuous variable. |
| Previous ectopic pregnancy | History of extrauterine pregnancy prior to the current gestation. | Continuous variable. |
| Threatened preterm labor | Regular uterine contractions with cervical changes (dilatation and effacement) between 23 and 36 weeks of gestation, with intact membranes. | Nominal variable. |
| Premature rupture of membranes (PROM) | Spontaneous rupture of membranes before the onset of labor, occurring prior to 37 weeks of gestation. | Nominal variable. |
| Intrauterine growth restriction (IUGR)<br>☐ Preterm<br>☐ At term | Fetal growth below the 10<sup>th</sup> percentile with Doppler abnormalities or estimated fetal weight below the 3<sup>rd</sup> percentile, either preterm (<37 weeks) or at term (≥37 weeks). | Nominal variable. |
| Preterm delivery | Gestational age less than 37 weeks at the time of | Nominal variable. |
|  | the end of the pregnancy. |  |
| Small-for-gestational age (SGA) | Birthweight below the 10<sup>th</sup> percentile but above the 3<sup>rd</sup> percentile for gestational age and sex, without Doppler abnormalities. | Nominal variable. |
| Pregnancy associated hypertension | Blood pressure $\geq 140/90$ mmHg identified after 20 weeks of gestation and resolving within 12 weeks postpartum. | Nominal variable. |
| Preeclampsia<br>☐ Preterm<br>☐ At term | Hypertension accompanied by proteinuria or signs of organ dysfunction (e.g., thrombocytopenia, renal/hepatic impairment, pulmonary edema, neurological symptoms). | Nominal variable. |
| Mode of delivery | Method by which the pregnancy ended (e.g., vaginal, instrumental, or cesarean delivery). | Nominal variable. |
| <u>Neonatal characteristics</u> |  |  |
| Neonatal weight | Baby's weight at birth. | Continuous variable, expressed in kilograms. |
| Apgar test score | Score measured at 1 and 5 minutes post-delivery to assess neonatal adaptation. | Continuous variable (0-10 score). |
| Cord blood pH | Measurement of pH in umbilical cord blood immediately after delivery | Continuous variable. |
| Intensive Care Unit (ICU) admission | Admission of the newborn to the Neonatal Intensive Care Unit (NICU), as determined by the attending pediatrician. | Nominal variable. |
| Neonatal morbidity | Any condition at birth potentially affecting neonatal adaptation or development. | Nominal variable. |

Data were extracted from electronic medical records after anonymization of sensitive information to ensure patient confidentiality. An effective screening and prevention strategy for preeclampsia was implemented, based on first-trimester assessments and ongoing monitoring throughout pregnancy.

### 2.3. Data processing and analysis

Data were analyzed using the Statistical Package for the Social Sciences (IBM SPSS Statistics, version 25.0). Nominal variables are presented as frequencies and percentages. Continuous variables are expressed as means with standard deviations (SD) or as medians with interquartile ranges (IQR), depending on the distribution of data. Normality was assessed using the Shapiro–Wilk test.

Categorical variables were compared using the Chi-square test or Fisher’s exact test, as appropriate. Continuous variables were analyzed using the student’s t-test. A p-value < 0.05 was considered statistically significant.

### 2.4. Ethical considerations

The study was approved by the Institutional Ethics Committee of Virgen del Rocío – Virgen Macarena University Hospitals (protocol number: 1154-N-23). Written informed consent was obtained from all participants prior to inclusion. Confidentiality was strictly maintained throughout the study in accordance with the principles of the Declaration of Helsinki. The data were used exclusively for research purposes.

## 3. Results

Between January and December 2024, a total of 128 pregnant women were enrolled in the study. All participants received comprehensive follow-up care in the Maternal-Fetal Medicine Unit of our hospital. Maternal demographic and obstetric characteristics are summarized in **Table 2 and Table 3**. The mean maternal age was 42 years (range: 40–50). Of the total sample, 104 women (81.25%) were aged between 40 and 45 years, while 24 women (18.75%) were aged 45 years or older. The mean BMI was 25.47 kg/m² (SD: 5.54), corresponding to the overweight category.

**Table 2.** Maternal demographic characteristics.

| Maternal demographic characteristics |  |
| --- | --- |
| Variables | Results |
| Maternal age, median (range) | 42 (40-50) |
| Weight, median (range) | 68.83 (44-120) |
| Height, n (%) | 164.55 (153-183) |
| BMI, median (range) | 25.47 (17.5-44.9) |
| Smoking status, n (%) | 12 (9.38%) |

**Table 3.** Obstetric characteristics.

| Obstetric characteristics |  |
| --- | --- |
| Variables | Results |
| Gravity, n (%) | 60 (46.88) |
| > n=1 | 68 (53.12) |
| > n=2 or more |  |
| History of miscarriage, n (%) | 52 (40.62%) |
| Parity, n (%) | 112 (87.5%) |
| > n=0 | 16 (12.5%) |
| > n=1 or more |  |
| Previous cesarean section, n (%) | 9 (7.03%) |
| Previous ectopic pregnancy, n (%) | 6 (4.68%) |

Approximately half of the participants were nulliparous (n = 60, 46.88%), and the vast majority were non-smokers (n = 116, 90.62%). A notable proportion had a history of miscarriage (n = 52, 40.62%), including 19 women with two or more previous spontaneous pregnancy losses. Additionally, six women (4.68%) reported a history of ectopic pregnancy. More than half of the participants (n = 71, 55.47%) conceived via ART using donated oocytes.

A remarkable proportion of women experienced preterm rupture of membranes (n = 11, 8.59%), diabetes mellitus (n = 6, 4.69%), or hypertensive disorders of pregnancy (n = 13, 10.32%), with seven of these cases presenting as preeclampsia (5.55%). The main obstetric outcomes are presented in **Figure 2**.

**Figure 2.**
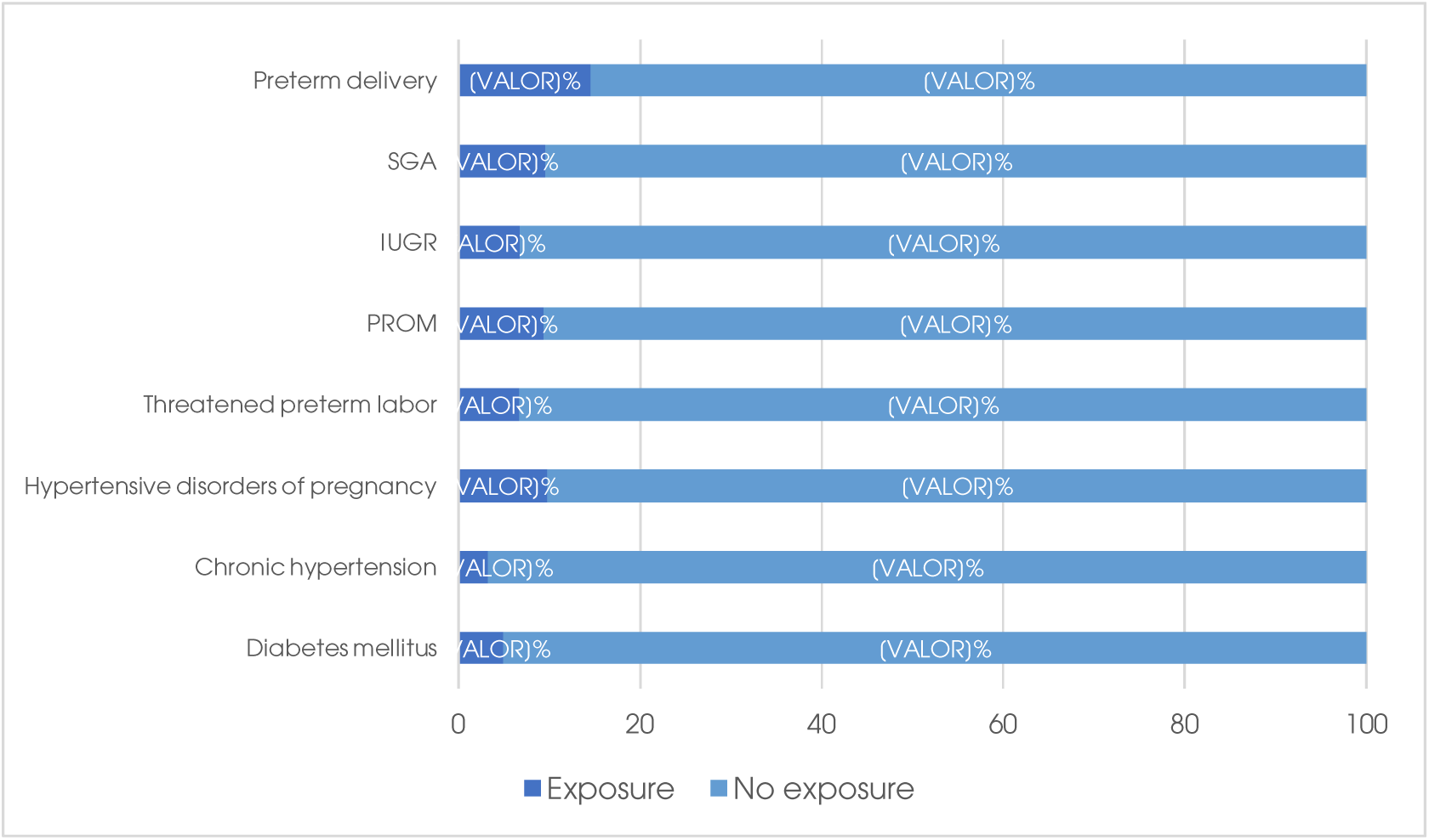
Principal obstetric outcomes.

The median gestational age at delivery was 37 weeks and 4 days (SD: 6.5). Cesarean section was performed in 63 cases (50.4%). The remaining pregnancies resulted in spontaneous vaginal delivery (n = 32, 25.6%) or operative vaginal delivery (n = 30, 24%).

Regarding neonatal outcomes, the mean birth weight was 3,027 grams (range: 840–4,340 g), and the mean umbilical cord pH was 7.29. Eight newborns (6.45%) required admission to the NICU. Neonatal comorbidities were identified in 14 infants, with respiratory complications being the most common, accounting for 12 of these cases. Neonatal outcomes are detailed in **Table 4**.

**Table 4.** Neonatal outcomes.

## 4. Discussion

This study examined the impact of AMA together with IVF on obstetric and perinatal outcomes. The women included in this cohort were closely monitored throughout pregnancy, allowing for early detection of complications. Nevertheless, despite careful surveillance, our findings indicate that women aged 40 years or older who conceived through IVF experienced higher rates of adverse perinatal outcomes compared to the general obstetric population.

Contrary to previous studies suggesting that appropriate prenatal follow-up and high-quality care can result in maternal and perinatal outcomes comparable to those of younger women^5^, our results revealed notably high rates of spontaneous miscarriage, diabetes mellitus, hypertensive disorders of pregnancy, preterm delivery, and cesarean section, among other relevant complications, in this population.

AMA is a well-established independent risk factor for first-trimester miscarriage, even after controlling for parity or a history of previous pregnancy loss^17^. The rate of first-trimester pregnancy loss ranges from 17% to 25% in women aged 35–40, increases to 33%–51% in those aged 40–45, and reaches 57%–75% in women over 45 years^11,17^. The proportion of women with a history of miscarriage in our study (40.62%) is consistent with these reported rates.

It has been observed that most pregnancy losses in AMA occur between 6 and 14 weeks of gestation^10^. These losses are often associated with chromosomal abnormalities such as trisomies and aneuploidies, likely resulting from diminished oocyte quality and functional impairments related to aging^10^.

In our analysis, the incidence of diabetes mellitus among AMA-IVF population was notably elevated (4.69%). It is well known that insulin resistance tends to increase with age, which in turn raises the risk of developing diabetes mellitus^3^. Prior studies have reported that the risk of both pre-existing and gestational diabetes is three to six times higher in women over 40 years of age compared to those aged 20–29^10^. In the general obstetric population, the incidence of gestational diabetes is approximately 3%, whereas it rises to 7%–12% in women over 40 and can reach up to 20% in those over 50^10^. Our findings align with this trend.

These results are further supported by data from the National Health and Nutrition Examination Survey, which showed that the prevalence of diabetes in non-pregnant women aged 35–39 was approximately two to three times higher, and three to five times higher in women aged 40–44, compared to those aged 20–34^1^.

From a biological standpoint, the association is plausible: reduced insulin sensitivity and progressive dysfunction of pancreatic β-cells—linked to endocrine aging—may contribute to the increased incidence of diabetes mellitus in women of AMA^3,11^.

AMA is also a well-documented risk factor for hypertensive disorders of pregnancy, particularly among women over 40 years of age^3,17,19^. Accordingly, it is reasonable to expect higher rates of chronic hypertension as well as an increased incidence of gestational hypertension and preeclampsia in this population^3^.

This assumption is confirmed by data obtained from general obstetric population. The incidence of preeclampsia is estimated at 3–4%, rising to 5–10% among women aged over 40, and up to 35% in those aged over 50^10^. This trend is reflected in our study, where 5.55% of pregnancies were complicated by preeclampsia.

This association is supported by both an earlier systematic review and a more recent meta-analysis focusing on women over the age of 40, which demonstrated a 1.5 to 2.0 times higher risk of developing preeclampsia in this population compared to their younger counterparts, whether they were primiparous or multiparous^17,20^. Several mechanisms have been proposed to explain this association, including reduced maternal hemodynamic adaptation during pregnancy, decreased compliance of uterine vasculature, and the presence of comorbidities such as elevated BMI, chronic hypertension, and preexisting diabetes mellitus^3,11^.

Another well-established concern associated with AMA is the increased rate of cesarean section. Numerous studies have demonstrated a linear relationship between advancing maternal age and the likelihood of cesarean delivery, with no clear threshold effect^17^. This pattern has been observed consistently across healthcare settings, although the magnitude of the association may vary depending on the population studied^17^.

For instance, a large cohort study including over 78,000 singleton births in the United States (2003–2012) revealed that the rate of primary cesarean section increased progressively with age, from 20% in women aged 25–34 years, to 26% in those aged 35–39, 31% in those 40–44, 36% in those 45–49, and as high as 61% in women over 50 years^10^. These findings are consistent with, yet lower than, the rate observed in our study, where 50.4% of the women underwent cesarean delivery.

However, we should note that the studies described above focus exclusively on healthy women with no fertility disorders or other concurrent pathologies, which implies considerable differences with our study sample. Addressing the role of ART is crucial given the described association between ART-treated pregnancies and higher rates of cesarean delivery^21,22^.

In Australia, a retrospective population-based study evaluated 17,019 women who underwent IVF and compared it with a national population of women who gave birth in the same country^23^. It was reported a rate of 50.1% for cesarean delivery in IVF-pregnancies versus 28.9% for all other births. In addition, the donor status was associated with significantly higher rates of cesarean section^23^.

Chien et al.^24^ have also noted this increased rate of elective cesarean delivery particularly among primiparous women with a singleton pregnancy who conceived following ART. After adjusting for sociodemographic characteristics such as maternal age over 35 years and other common comorbidities, ART-pregnancies were 2.95 times more likely to end in planned cesarean deliveries^24^.

The reasons behind the increasing rate of cesarean section in AMA remain debated^17^. One hypothesis is that the aging uterus may be less efficient at generating effective contractions, contributing to higher rates of labor dystocia. However, in vitro studies on myometrial contractility have shown mixed results^3,17^. Additional factors such as atherosclerotic changes in uterine arteries, reduced oxytocin receptor expression, and impaired myometrial function have also been implicated^7,9^.

Moreover, increased awareness among both healthcare providers and patients of the heightened cesarean risk in AMA and IVF may lower the threshold for surgical intervention^17,25^. Finally, both conditions are frequently associated with complications such as hypertensive disorders and placental abnormalities, for which cesarean delivery may be indicated on maternal or fetal grounds^12,22^.

In possible association with the elevated cesarean section rate observed in our study, the incidence of preterm delivery is also notable. A significant proportion of women of AMA experienced preterm birth (12.7%), which is clearly higher than the rate reported in the general obstetric population (4.1%)^16^.

A cohort study involving 173,715 healthy nulliparous women with singleton pregnancies demonstrated that maternal age between 35 and 40 years was significantly associated with an increased risk of preterm birth, even after adjusting for potential confounding factors such as smoking, parity, and pre-existing maternal diseases^10^. Similarly, a systematic review and meta-analysis conducted by Pinheiro et al. comparing obstetric and perinatal outcomes in singleton pregnancies among AMA women (35–40 and >40 years old) versus younger women (20–34 years) confirmed that AMA was associated with a higher rate of preterm delivery^5^.

The mechanisms underlying this association remain insufficiently understood^3,17^. A Canadian study on biological determinants of preterm birth indicated that hypoxic conditions, such as those caused by hypertensive disorders, contribute to preterm birth risk by reducing placental perfusion and limiting the supply of oxygen and nutrients to the fetus^1^. Additionally, iatrogenic factors—such as the need for early induction of labor due to maternal or fetal complications—may also play a role^3^. It is likely that age-related physiological changes contribute to this association, although these mechanisms remain to be fully elucidated^4^.

As a potential consequence of the elevated preterm birth rate in our cohort, 6.45% of newborns required admission to the NICU. Previous research has shown that NICU admission rates increase progressively with maternal age, peaking at approximately 6% among women aged 45–46 years^4^. Likewise, Pinheiro et al. reported a higher risk of NICU admission among neonates born to AMA mothers (age >40, OR 1.20; 95% CI: 1.13–1.27; I² = 0%) compared to those born to women aged 35–40 years (OR 1.13; 95% CI: 1.09–1.18; I² = 47%)^5^.

Moreover, a study by Newman et al. examining the effects of IVF in women aged 45 years or older found that neonates conceived through IVF were significantly more likely to require NICU admission and had a longer length of hospital stay (>4 days)^26^. These findings suggest that AMA and IVF may act cumulatively to increase the risk of adverse neonatal outcomes such as NICU admission.

The findings outlined above suggest that IVF pregnancies in women of AMA would benefit from the development of targeted awareness programs, thereby supporting couples throughout all stages of pregnancy. In particular, we propose a closer, comprehensive, and personalized monitoring through medical reviews at least once a month in order to anticipate the onset of prevalent obstetric complications, such as preeclampsia and diabetes, among others.

Above all else, our goal is contributed to improving the quality of perinatal care for older mothers. Evidence-based interventional strategies may also help to prevent adverse perinatal outcomes in this growing population.

## Strengths and limitations

Our study has several notable strengths. Chief among these is the comprehensive examination of the risk for multiple adverse obstetric and perinatal outcomes in a cohort of women of AMA undergoing IVF. To our knowledge, this represents the largest analysis to date focusing on this specific subgroup, addressing an important gap in the literature. Consequently, our findings may serve as baseline evidence, particularly in regions where no previous studies have been conducted. Additionally, the results can provide valuable guidance for healthcare professionals and AMA mothers regarding expected perinatal outcomes. This could also serve as a basis for the development and implementation of new clinical practice guidelines at the national or even international level.

Furthermore, the rigorous and prospective data collection methodology ensures the accuracy and reliability of the information while minimizing recall bias. The study was also strengthened by the use of a standardized maternal care protocol consistently applied throughout the study period. This approach promotes equity and consistency in prenatal care and reinforces the thoroughness of data capture.

Nevertheless, several limitations should be considered when interpreting our results. First, the single-center design may limit the generalizability of our findings, as the characteristics of AMA women in our hospital may differ from those in other settings.

The relatively small sample size, along with the limited study period and single-center scope, may reduce the power to detect statistically significant differences. Moreover, there may be unmeasured confounding factors not captured in our data that could bias and affect the precision of our estimates.

## 5. Conclusion

Pregnancies in women of AMA conceived via in IVF are associated with significant obstetric and neonatal risks. It is essential that both healthcare providers and patients are fully informed about these risks.

Our findings highlight that, even with meticulous, individualized monitoring in a maternal-fetal medicine unit specialized in high-risk pregnancies, not all adverse outcomes can be prevented.

Further research is warranted to better delineate the specific contributions of AMA and IVF to these complications. Clear management guidelines for AMA patients undergoing IVF should be developed in the future.

## Supporting information

Title page

## Data Availability

The datasets generated and analyzed during the current study are available from the corresponding author on reasonable request.

## Acknowledgements

All persons that contributed to this study are listed authors and meet the criteria for authorship.

## Disclosures

### Human rights statements and informed consent

All procedures followed were in accordance with the ethical standards of the responsible committee on human experimentation and with the Helsinki Declaration of 1964 and its later amendments. Informed consent was obtained from all patients for being included in the study.

### Animal studies

Not applicable.

### Approval by Ethics Committee

The research project including human subjects has been approved by a suitably constituted Ethics Committee (Ethics Committee of Virgen del Rocío – Virgen Macarena University Hospitals).

### Clinical Trial Registry

Not applicable.

### Authoŕs contribution

All listed authors have contributed to the manuscript substantially and have agreed to the final submitted version. M.D., B.W., J.D., A.C., G.A., and L.G. contribute to conception and design. M.D. and B.W. were responsible to acquisition of data. M.D., B.W., J.D., A.C., G.A., and L.G. contributed to the analysis and interpretation of data. All authors contribute to drafting the article or revising it critically for important intellectual content. All authors read and approved the final manuscript.

## List of abbreviations

ART: Assisted Reproductive Technologies
IVF: In Vitro Fertilization
AMA: Advanced Maternal Age
NICU: Neonatal Intensive Care Unit
BMI: Body Mass Index

